# Examining the association between family status and depression in the UK Biobank

**DOI:** 10.1101/2020.07.07.20148023

**Authors:** Alexandros Giannelis, Alish Palmos, Saskia P. Hagenaars, Gerome Breen, Cathryn M. Lewis, Julian Mutz

**Author notes:** **Corresponding author:** Julian Mutz; Social, Genetic & Developmental Psychiatry Centre, Institute of Psychiatry, Psychology & Neuroscience, King’s College London; Memory Lane, London SE5 8AF, United Kingdom.

## Abstract

**Background:** We examined associations between family status (living with a spouse or partner, number of children) and lifetime depression.

**Methods:** We used data from the UK Biobank, a large prospective study of middle-aged and older adults. Lifetime depression was assessed as part of a follow-up mental health questionnaire. Logistic regression was used to estimate associations between family status and depression. We included extensive adjustment for social, demographic and other potential confounders, including depression polygenic risk scores.

**Results:** 52,078 participants (mean age=63.6, SD=7.6; 52% female) were included in our analyses. Living with a spouse or partner was associated with substantially lower odds of lifetime depression (OR=0.67, 95%CI 0.62-0.74). Compared to individuals without children, we found higher odds of lifetime depression for parents of one child (OR=1.17, 95%CI 1.07-1.27), and parents of three (OR=1.11, 95%CI 1.03-1.20) or four or more children (OR=1.27, 95%CI 1.14-1.42). Amongst those not cohabiting, having any number of children was associated with higher odds of lifetime depression. Our results were consistent across age groups, the sexes, neighbourhood deprivation and genetic risk for depression. Exploratory Mendelian randomisation analyses suggested a causal effect of number of children on lifetime depression.

**Limitations:** Our data did not allow distinguishing between non-marital and marital cohabitation. Results may not generalise to all ages or populations.

**Conclusions:** Living with a spouse or partner was strongly associated with reduced odds of depression. Having one or three or more children was associated with increased odds of depression, especially in individuals not living with a spouse or partner.

## Introduction

Major depressive disorder currently affects up to 322 million people worldwide, making it the most prevalent mood disorder (1). At least half of the global population report experiencing depressive symptoms at some point in their lives (2). Up to a third of older adults suffer from depression, many of whom experience functional decline and cognitive impairment (3). Depression is the fifth leading cause of long-standing disability amongst all diseases, and the first amongst mental health disorders (4). It has been associated with lower productivity (5,6), lower earnings (7,8), increased risk of suicide (9,10) as well as higher rates of cardiovascular disease (11), cancer (12) and all-cause mortality (13).

Family status, an umbrella term which includes marital status, cohabiting with a spouse or partner and parenthood, has consistently been associated with the occurrence and severity of depression (2,14). For example, depression has been associated with lower odds of ever getting married (odds ratios ranging from 0.6 to 0.8) (15–17). A longitudinal study found an 11% reduction in the probability of getting married for children reporting depressive symptoms, compared to their siblings (18). Large meta-analyses of studies in Chinese older adults have shown that the prevalence of depression is substantially higher amongst non-married (including divorced, unmarried and widowed) individuals (19,20). International data follow a similar pattern (21), with married individuals reporting higher levels of well-being across cultures (22). In women, not being married is associated with a higher incidence of postnatal depression (23). Married individuals tend to be healthier, happier and less depressed (24). Based on these findings it has been suggested that marriage is a protective factor which shields individuals from stressful circumstances and provides confidant support (25). Findings from twin studies which attempt to control for genetic predisposition to depression have shown that divorced, widowed and never-married twins had higher rates of depression, indicating a protective effect of marriage (26–28). Similar associations have been found for physical health, with married individuals having a lower risk of cardiovascular disease (29,30). Concerning non-marital cohabitation, data from the Health and Retirement Study suggested that non-married cohabiting individuals had a higher risk of depression compared to married individuals, but lower compared to never-married, divorced or widowed individuals (31).

Parenthood has been associated with lower rates of depression, with one study showing that depressive symptoms decrease as the number of children increases (32). Another study reported no association between depression and parenthood in general, but positive associations with certain types of parenthood (for example, in single parents and parents of young children) (33). However, these associations might be modified by marital status – with single parents reporting more depressive symptoms – as well as by cultural and ethnic factors (33,34). Single mothers in particular suffer from high rates of depression and it has been suggested that this association is mediated by high levels of stress and lack of social support (35,36). The association might also be confounded by postnatal depression which affects up to 15% of mothers (37).

There are a number of factors associated with both family status and depression, thus making it challenging to dissect associations and causal pathways. These factors include socioeconomic and demographic characteristics, personality traits and life events. See Supplement 1 for an overview of potential confounders. In addition to environmental factors, genetic predispositions should be considered in epidemiological research on depression. Twin and adoption studies have suggested that depression is moderately heritable, with approximately 37% of the variance in depression status being due to additive genetic effects (38). Recent genome-wide association studies (GWAS) that test millions of single nucleotide polymorphisms (SNPs) across the genome have identified >100 SNPs associated with depression (39,40). Approximately 10% of the variance in depression status can be explained by common SNPs. GWAS summary statistics can be used to create individual-level polygenic risk scores (PRS), which provide a measure of genetic liability for the relevant trait. Although the amount of variance in depression explained using PRS is small, such scores might be useful for risk stratification. For example, individuals in the highest depression PRS decile were 2.5 times more likely to be depressed, compared to those in the lowest decile (39).

Previous studies of the association between family status and depression had various limitations, mainly failing to control for potential confounders. For example, the higher level of income and education of married individuals may be one reason for the lower prevalence of depression amongst them. Another possibility is that people who are less genetically inclined to become depressed may be those who enter partnerships and have children more frequently. The present study contributes to the literature by using a large sample and controlling for a multitude of non-genetic factors as well as a PRS for depression.

The aim of the present study was to examine the association between family status (i.e. living together with a spouse or partner; and number of children) and depression in the UK Biobank. Based on previous research (26,30) we hypothesised that living with a partner or spouse is associated with lower odds of depression. We also aimed to explore the association between the number of children and depression for which the evidence has so far been inconsistent (32,33). An additional research aim was to discern if associations between family status and depression differed by sex, age group, neighbourhood deprivation or PRS for depression.

## Methods

### Study description

The UK Biobank is a prospective study of approximately 500 000 men and women, aged 37-73, recruited between 2006-2010. Individuals who were living within a 25-mile (∼40 km) radius of one of 22 assessment centres across England, Scotland and Wales were invited to participate. At the baseline assessment, participants provided informed consent and reported sociodemographic, lifestyle and medical history factors through touch-screen questionnaires and nurse-led interviews. They also provided biological samples (blood, urine and saliva), physical measurements (e.g. height and weight) and consented to their data being linked to their health records. The study has been described in detail in previous publications (41,42).

The present study is based on participants who completed an online follow-up mental health questionnaire (MHQ) between 2016-2017. The questionnaire was developed by a team of experts and includes information on lifetime depression assessed using the Composite International Diagnostic Interview Short Form (CIDI-SF) (43). Almost 340 000 participants were invited via e-mail to complete a “thoughts and feelings questionnaire” and more than 150 000 participants have provided responses (44).

### Family status

Two variables were included for family status: cohabitation status and number of children. Participants who answered “Husband, wife or partner” in response to the question “How are the other people who live with you related to you?” were classified as “living with a spouse or partner”. The number of biological children was based on “Number of live births” for women and “Number of children fathered” for men. We categorised participants into five groups: “childless”, “parent of one”, “parent of two”, “parent of three” and “parent of four or more”.

### Depression

Lifetime depression was assessed using the depression module of the CIDI-SF and defined according to DSM-5 criteria for major depressive disorder. Lifetime severe depression was defined as meeting criteria for lifetime depression, endorsing all non-core symptoms on the CIDI-SF and reporting a lot of impairment in normal functioning. Current depression was defined as meeting criteria for lifetime depression and reporting at least five symptoms, including at least one core symptom, on the Patient Health Questionnaire 9 (PHQ-9) more than half the days (several days for recent thoughts of suicide or self-harm) over the past two weeks (45,46). Current severe depression was defined as meeting criteria for current depression and having a sum score of 15 or more on the PHQ-9. See Supplement 2 and (44).

### Covariates

Potential confounders of the association between family status and depression were identified from the baseline assessment data (sex, marital separation/divorce in the two years prior to assessment, death of a spouse/partner in the two years prior to assessment, migrant status, highest educational or professional qualification, annual gross household income, employment status, Townsend deprivation index, smoking status, long-standing illness, disability or infirmity, neuroticism, participation in leisure/social activities, loneliness, ever had same-sex intercourse, lifetime number of sexual partners and body mass index) and from the MHQ (age at completing the questionnaire, alcohol use, adverse childhood experiences and traumatic life events) (Supplement 3). We also included a PRS based on summary statistics from the most recent depression GWAS, excluding UK Biobank participants (39) (see Supplement 4).

### Exclusion criteria

Individuals with missing data or who responded “prefer not to answer” or “do not know” were excluded from all analyses. The lifetime number of sexual partners was truncated at the 99th%ile (> 50). We also excluded individuals with self-reported psychosis or mania on the MHQ. Of the individuals who did not meet criteria for depression as described above, we excluded any participant who had self-reported any other mental disorder, if they were currently taking antidepressant medication (Supplement 5), if their Hospital Episode Statistics record contained any mood disorder diagnosis (Supplement 6) or if they were classified as individuals with bipolar or major depressive disorder according to Smith et al. (47) (Supplement 7). See also Supplement 2.

### Statistical analysis

Our main analysis focused on lifetime depression (i.e. having ever met diagnostic criteria for major depressive disorder). We used logistic regression to estimate the association between family status and depression. For each of the two explanatory variables (cohabitation status and number of children) we first fitted crude models without adjustment for potential confounders. In subsequent models we (i) adjusted for age and sex and (ii) adjusted for age, sex, all other non-genetic covariates, the depression PRS, six ancestry-informative principal components as well as batch and assessment centre. We also fitted an additional model that included both cohabitation status and number of children as well as all covariates (termed the “fully adjusted model”).

Multicollinearity was assessed using generalised variance inflation factors (48). For all reported *p*-values, multiple testing correction was performed using the Benjamini & Hochberg false discovery rate approach (49). For each explanatory variable, we calculated its statistical significance adjusted for a 5% false discovery rate, by taking into account the variable’s *p*-values from the four main regressions and the four sensitivity analyses (further details below). Statistical significance for cohabitation status was thus corrected for eight tests, while statistical significance for number of children was corrected for 32 tests (eight regression models × four levels of the explanatory variable).

We performed additional analyses of the fully adjusted model that included both explanatory variables and covariates, stratified by age group (decades), sex, Townsend deprivation index quintile and depression PRS quintile to investigate whether any associations between family status and depression differed by these characteristics. We also performed an analysis of the model that included number of children and covariates, stratified by cohabitation status to assess associations between single parenthood and depression. Finally, we added the following cross-product terms individually to the fully adjusted model to test for interaction effects: cohabitation status × age, cohabitation status × sex, cohabitation status × Townsend deprivation index, cohabitation status × depression PRS, cohabitation status × number of children, number of children × age, number of children × sex, number of children × Townsend deprivation index, and number of children × depression PRS.

### Sensitivity analysis

To assess whether our findings were consistent across different depression phenotypes, we substituted current depression for lifetime depression in the fully adjusted model. We also restricted analyses to individuals with severe depression for both lifetime and current depression (Supplement 2).

Finally, we repeated the main analysis after excluding individuals with postnatal depression which might confound the association between family status and depression. The association between depression and childbirth, which is associated with hormonal changes and circumstances during pregnancy and birth (50), should be considered separately from that of parenthood in general. Since we were interested in the effect of parenthood and not childbirth per se, we excluded women who reported depression that was possibly related to childbirth.

All phenotypic analyses were performed in R (version 3.5.0).

### Mendelian randomisation

Exploratory mendelian randomisation (MR) analyses were performed to test for any causal genetic associations between lifetime depression and family status. Our primary MR analyses were carried out using Generalised Summary-data-based Mendelian Randomisation (GSMR) (51). The GSMR method tests for putative causal association between a risk factor and an outcome using summary-level data from GWAS analyses (see Supplement 8) and requires a reference sample with individual level genotypes for linkage disequilibrium (LD) estimation. In the present study we used the 1000 Genomes Project as reference sample, with a clumping r^2^ threshold of 0.05 and an FDR threshold (to shrink chance correlations between SNP instruments to zero) of 0.05 (52). The HEIDI-outlier method threshold for detecting pleiotropy was set to 0.01. This is a robust method for detecting and eliminating genetic instruments that have pleiotropic effects on both the exposure and outcome (51).

Due to the small number of SNPs significant above the standard *p* < 5 × 10^−8^ threshold, we lowered the threshold in the present study to *p* < 5 × 10^−7^ for lifetime depression and number of children, and *p* < 5 × 10^−6^ for cohabitation status. This resulted in 12, 21 and 18 SNPs that could be used as genetic instruments, respectively. We performed bi-directional MR analyses between number of children and lifetime depression, and between cohabitation status and lifetime depression.

To validate the findings from GSMR, a secondary MR analysis was carried out using the Two-Sample MR package in R (53). The same genetic instruments were used for each GWAS as for GSMR analyses. The same bi-directional analyses were performed using the Inverse Variance Weighted (IVW), MR-RAPS and MR-Egger methods (54). These have been described as robust MR methods and are commonly used as sensitivity analyses when performing MR (54,55).

## Results

### Descriptive statistics

Of the 157 389 MHQ respondents, 126 315 were retained after genetic quality control (see Supplement 4). After excluding individuals with missing data on depression, family status or covariates, 52 078 participants were included in our main analysis (Figure 1). The average time between the baseline assessment and completion of the MHQ was 8.11 years (SD = 0.88).

**Figure 1.**
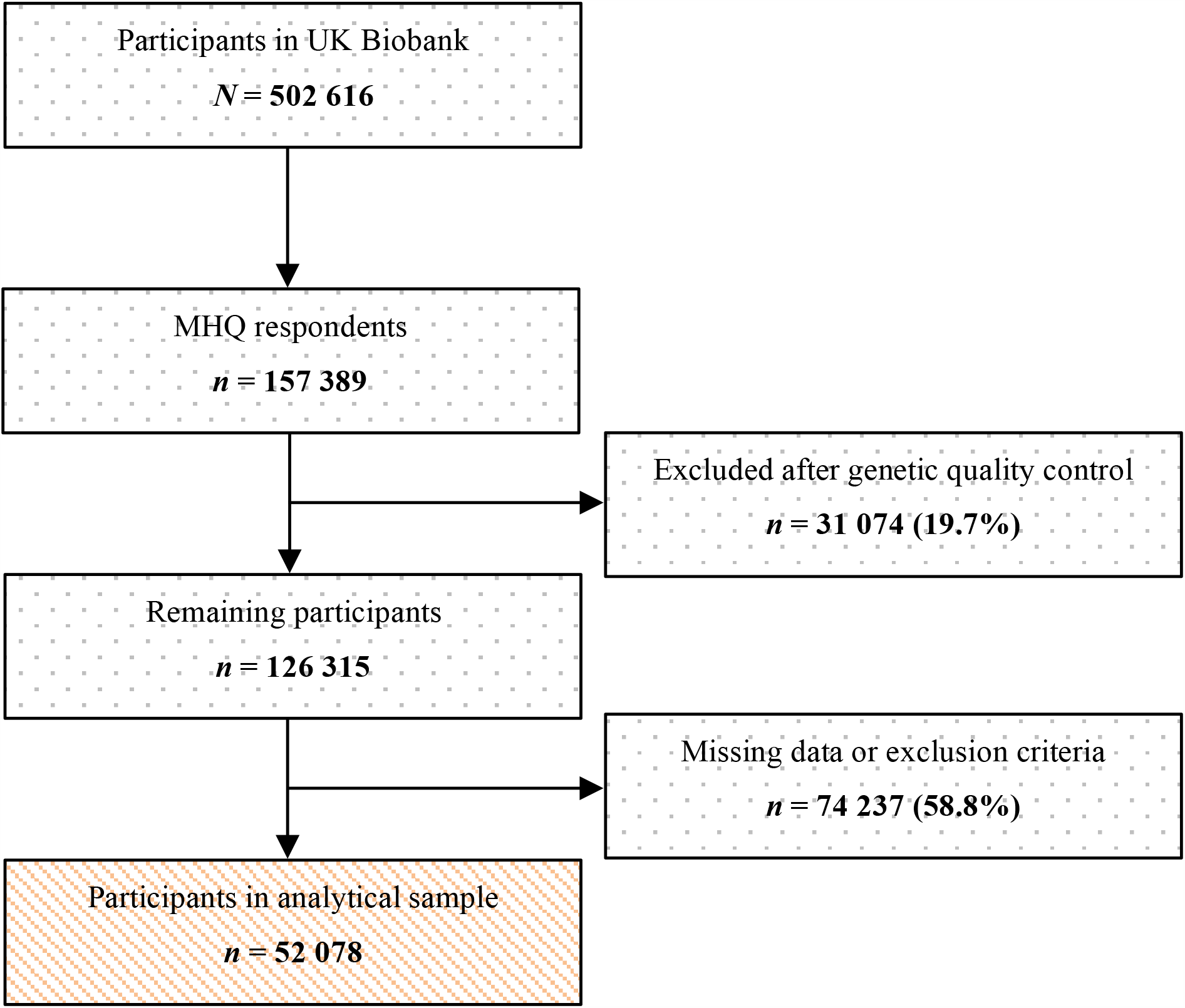
Flowchart of study sample.

Table 1 illustrates the characteristics of the full and analytical sample. The percentage of individuals in the analytical sample who were cohabiting with a spouse or partner was higher than that of the UK general population (93% compared to 69% of those aged 45 and above) (56). There were notable differences in rates of lifetime depression between individuals who reported cohabiting with a spouse or partner and those who did not (27% and 50%, respectively) (Figure 2). However, lifetime depression rates were fairly consistent across parental categories, ranging from 27% for parents of two children to 35% for parents of one child (Figure 2). The mean age at first episode of depression was 37.3 years (SD = 14.73), while the mean age at the most recent depressive episode was 48.9 years (SD = 12.79).

**Table 1.**
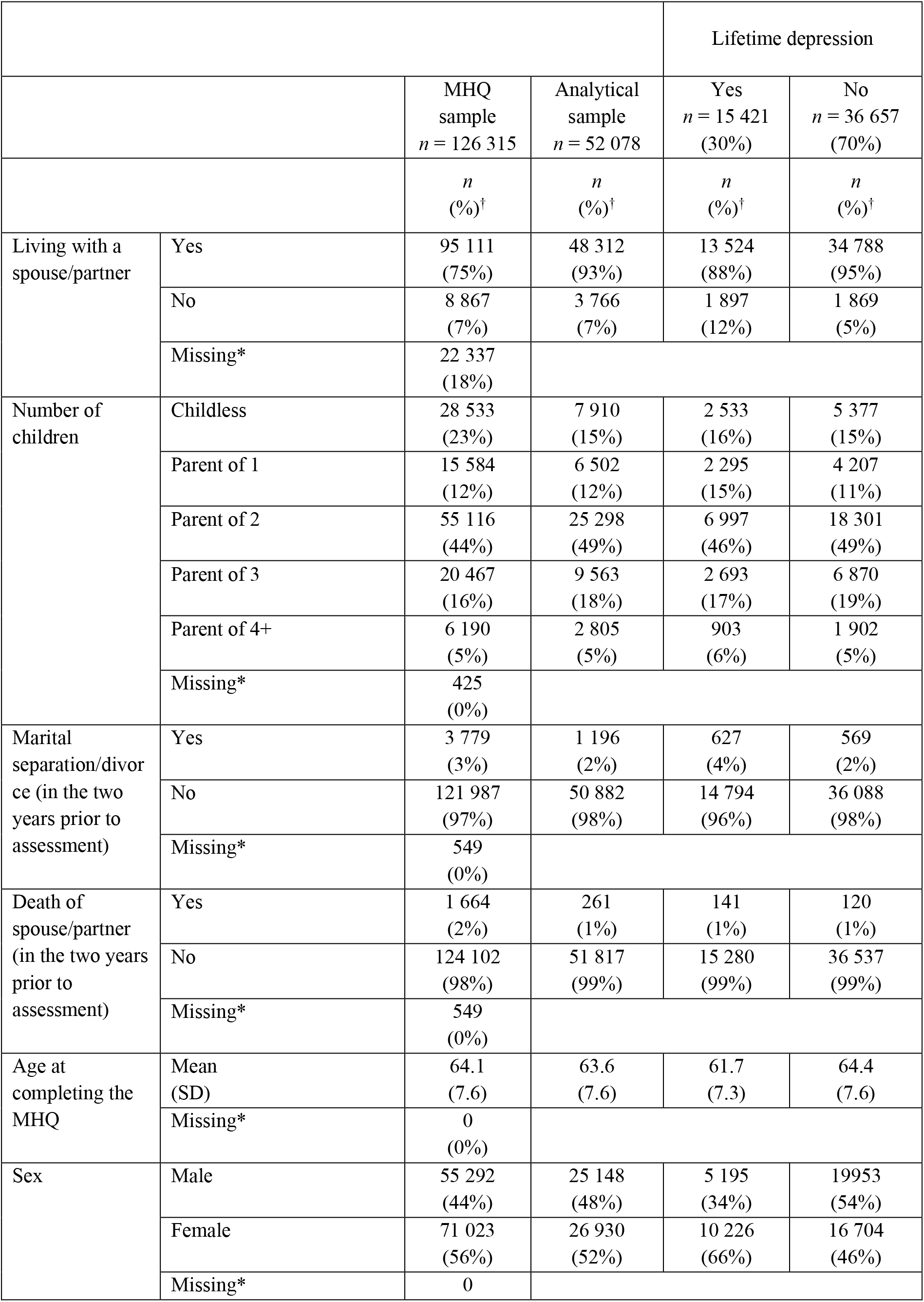
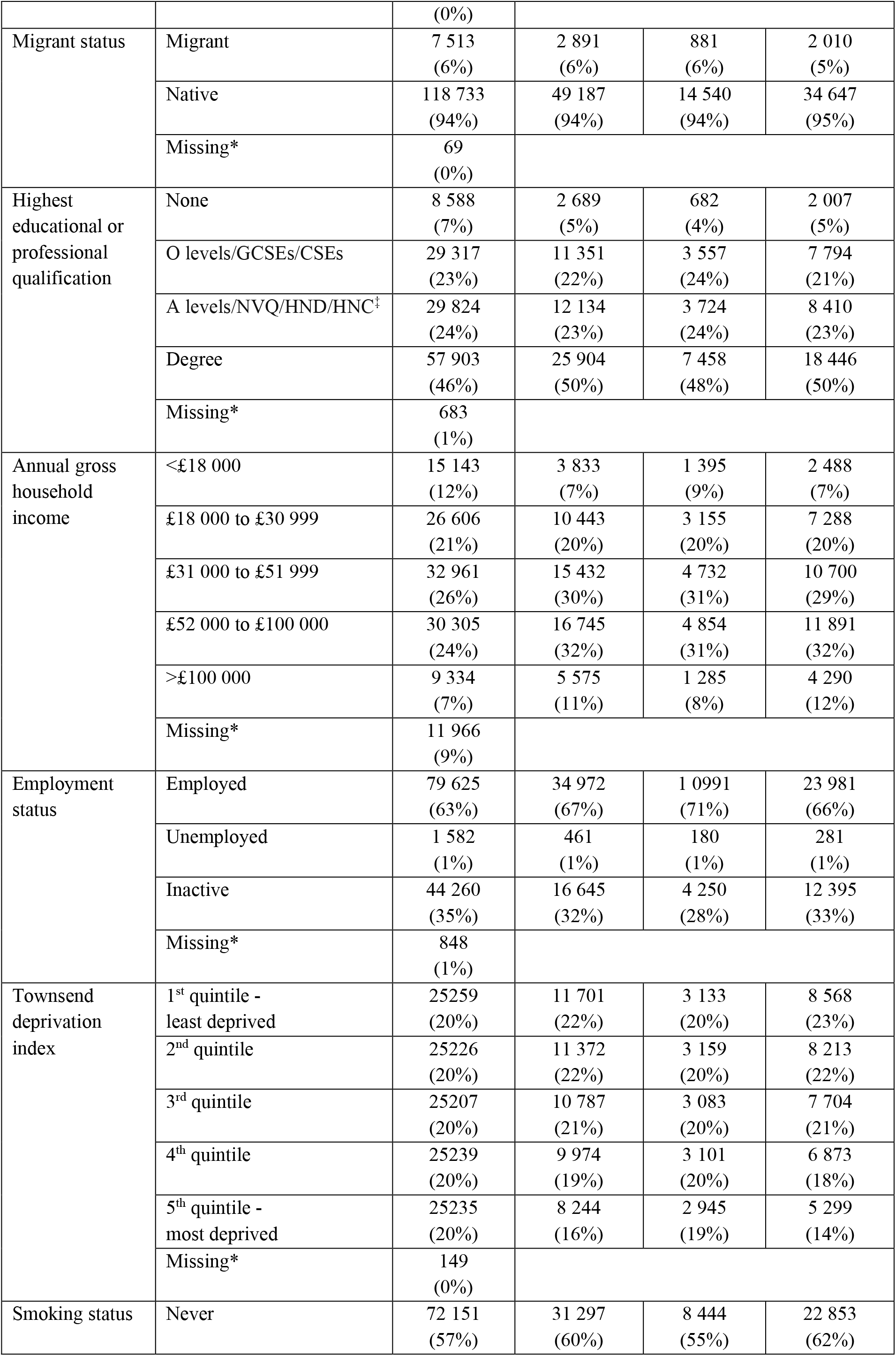
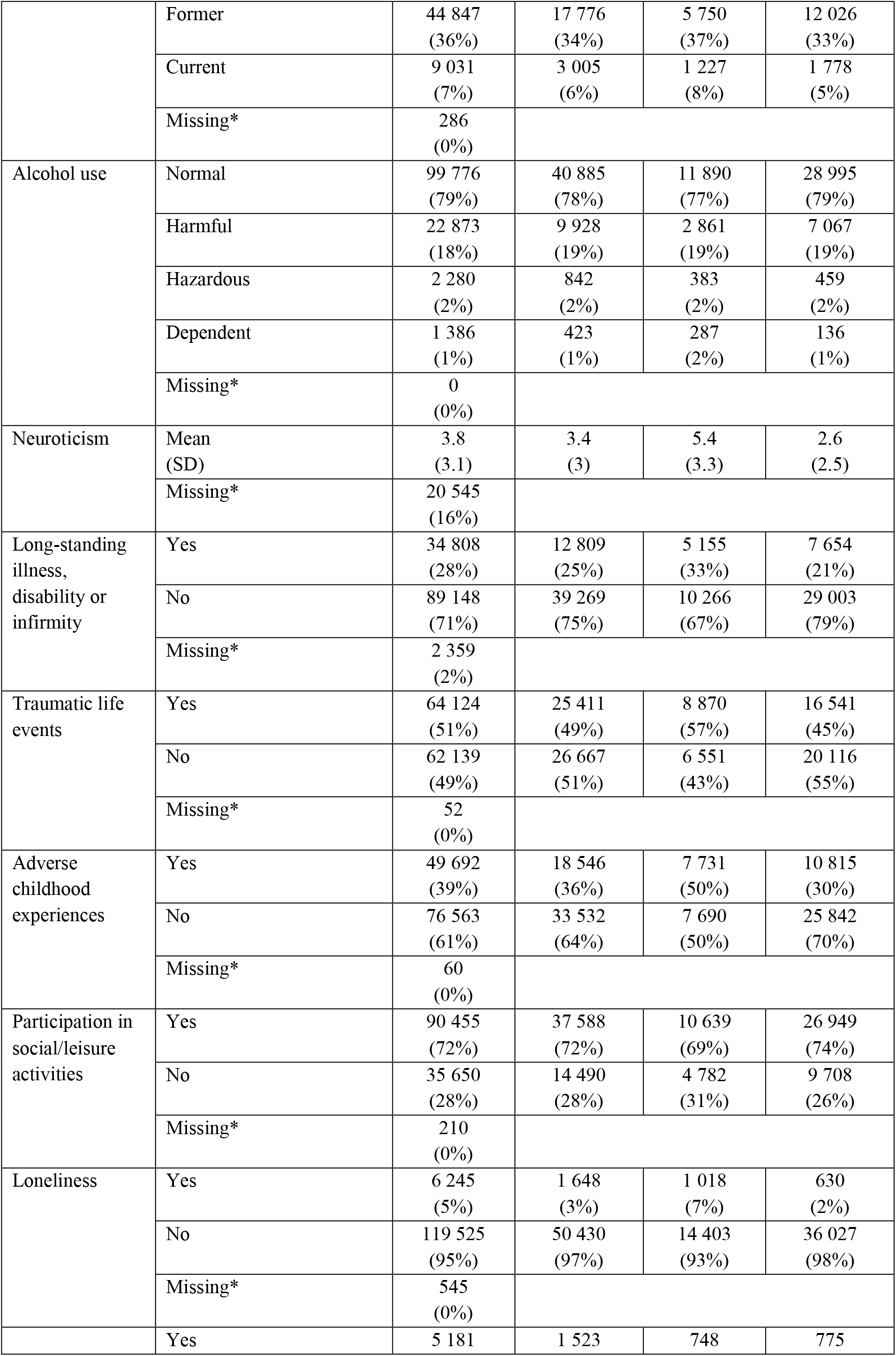
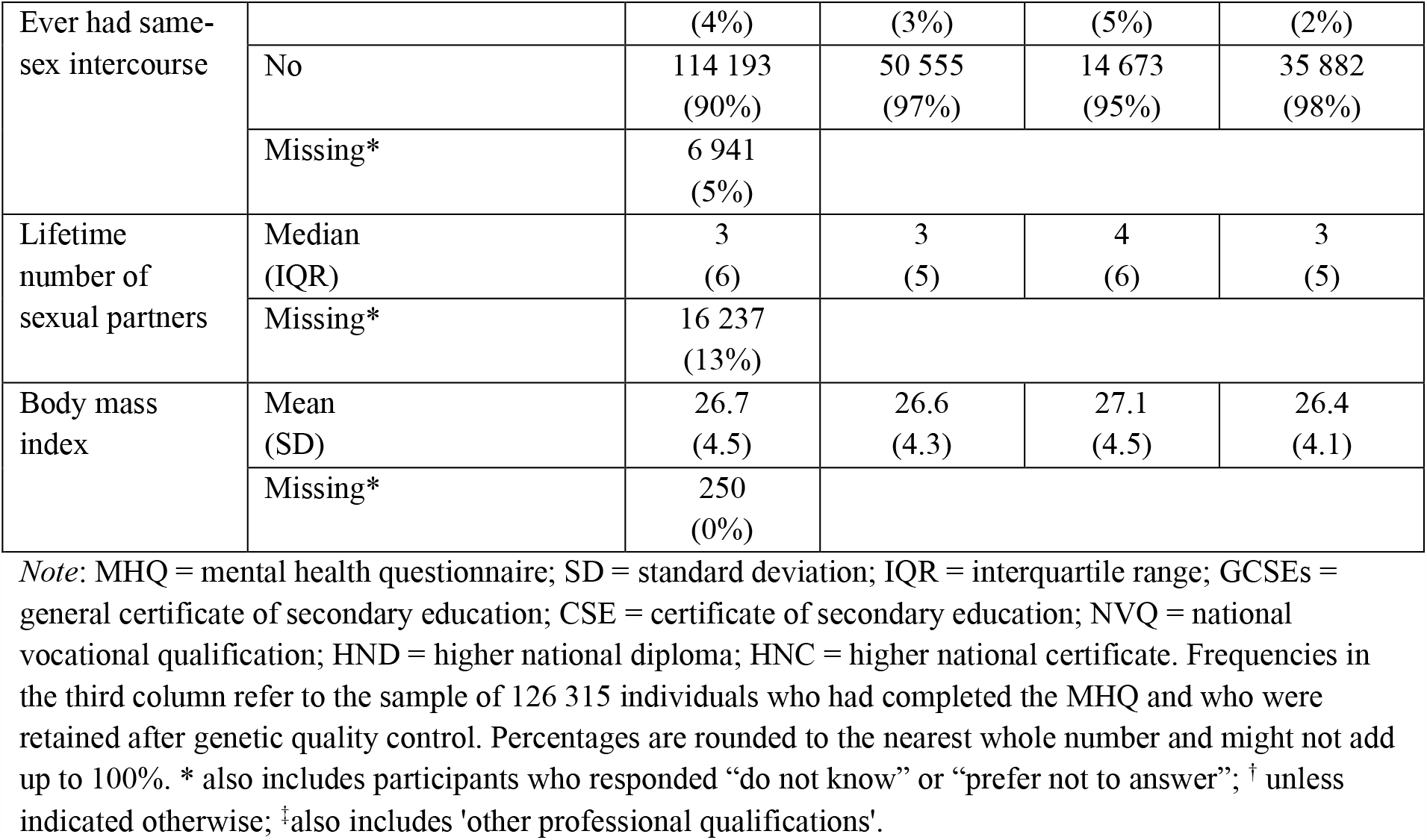
Sample characteristics

**Figure 2.**
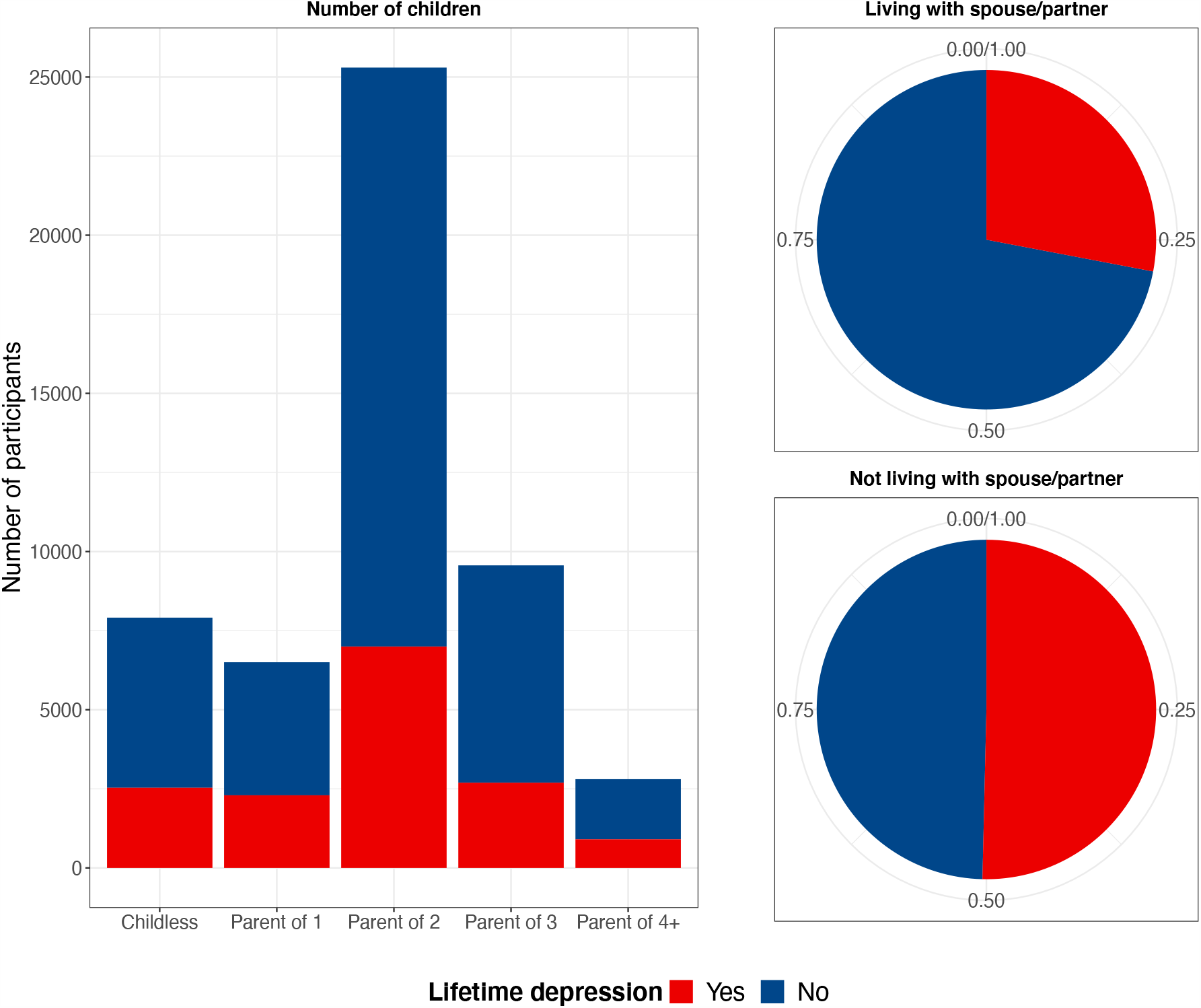
Lifetime depression by number of children and cohabitation status.

### Main analysis

Table 2 presents the results of the logistic regression analyses with lifetime depression (yes/no) as outcome. Generalised variance inflation factors were all less than 2.56, indicating low levels of collinearity between explanatory variables included in the fully adjusted model.

**Table 2.** Association between family status and lifetime depression

| *n* = 52 078 | Unadjusted | Adjusted for age and sex | Adjusted for all covariates<sup>†</sup> | Fully adjusted model with both family status variables |
| --- | --- | --- | --- | --- |
| **Cohabitation status** (living with spouse/partner) |  |  |  |  |
| No | Ref | Ref | Ref | Ref |
| Yes | 0.38***<br>(0.36, 0.41) | 0.50***<br>(0.47, 0.53) | 0.67***<br>(0.61, 0.73) | 0.67***<br>(0.62, 0.74) |
| **Number of children** |  |  |  |  |
| Childless | Ref | Ref | Ref | Ref |
| Parent of 1 | 1.16***<br>(1.08, 1.24) | 1.21***<br>(1.12, 1.29) | 1.19***<br>(1.10, 1.29) | 1.17***<br>(1.07, 1.27) |
| Parent of 2 | 0.81***<br>(0.76, 0.85) | 0.91***<br>(0.85, 0.95) | 1.02<br>(0.95, 1.09) | 1.01<br>(0.95, 1.08) |
| Parent of 3 | 0.83***<br>(0.77, 0.88) | 0.96<br>(0.91, 1.03) | 1.12**<br>(1.04, 1.21) | 1.11*<br>(1.03, 1.20) |
| Parent of 4+ | 1.00<br>(0.91, 1.10) | 1.25***<br>(1.13, 1.37) | 1.28***<br>(1.14, 1.43) | 1.27***<br>(1.14, 1.42) |
| *Note:* <sup>†</sup> adjusted for age, sex, marital separation/divorce (in the two years prior to assessment), death of spouse/partner (in the two years prior to assessment), migrant status, highest educational/professional qualification, annual gross household income, employment status, Townsend deprivation index, smoking status, alcohol use, neuroticism, long-standing illness, disability or infirmity, traumatic life events, adverse childhood experiences, participation in social/leisure activities, loneliness, ever had same-sex intercourse, lifetime number of sexual partners, body mass index, depression polygenic risk score, six ancestry-informative principal components, batch number and assessment centre. *** *p* < 0.001, ** *p* < 0.01, * *p* < 0.05 (corrected for multiple testing at 5% false discovery rate). |  |  |  |  |

Cohabitation status was associated with lifetime depression across models, with 33% to 62% lower odds of lifetime depression in participants living with a spouse or partner. Adjusting for potential confounders and number of children attenuated these associations (ranging from OR = 0.38, 95% CI 0.35-0.41 in the unadjusted model to OR = 0.67, 95% CI 0.62-0.74 in the fully adjusted model).

Findings regarding the association between number of children and lifetime depression were mixed. Compared to individuals without children, parents of one child had higher odds of lifetime depression across models (OR = 1.17, 95% CI 1.07-1.27 in the fully adjusted model). While parents of two children had lower odds of lifetime depression in the unadjusted model (OR = 0.81, 95% CI 0.76-0.85) and after adjustment for age and sex (OR = 0.91, 95% CI 0.85-0.95), we did not find evidence of an association with lifetime depression after controlling for other confounders and cohabitation status. Parents of three children or four or more children had higher odds of lifetime depression in the fully adjusted model (OR = 1.11, 95% CI 1.03-1.20 and OR = 1.27, 95% CI 1.14-1.42, respectively), compared to individuals without children.

### Stratified analysis

Associations between cohabitation status and lifetime depression were of similar magnitude and direction across age groups (Figure 3), and there was no evidence of an interaction between cohabitation status and age in the full analytical sample (*p*_interaction_ = 0.44). For participants aged 45-54 (*n* = 8 108) and 75-80 (*n* = 3 264) years, there was no evidence of an association between number of children and lifetime depression. We also did not find evidence of an interaction between number of children and age (all *p*_interaction_ > 0.05). Associations between cohabitation status and lifetime depression were similar for both sexes (Figure 4), and there was no evidence of an interaction between cohabitation status and sex (*p*_interaction_ = 0.93). There was also no evidence of an interaction between number of children and sex (all *p*_interaction_ > 0.05). Associations between family status and lifetime depression were fairly consistent across depression PRS quintiles (Figure 5), and there was no evidence of an interaction between family status and the depression PRS (all *p*_interaction_ > 0.22).Associations between family status and lifetime depression were also fairly consistent across Townsend deprivation index quintiles (Figure 6), and there was no evidence of an interaction between family status and Townsend deprivation index (all *p*_interaction_ > 0.19).

**Figure 3.**
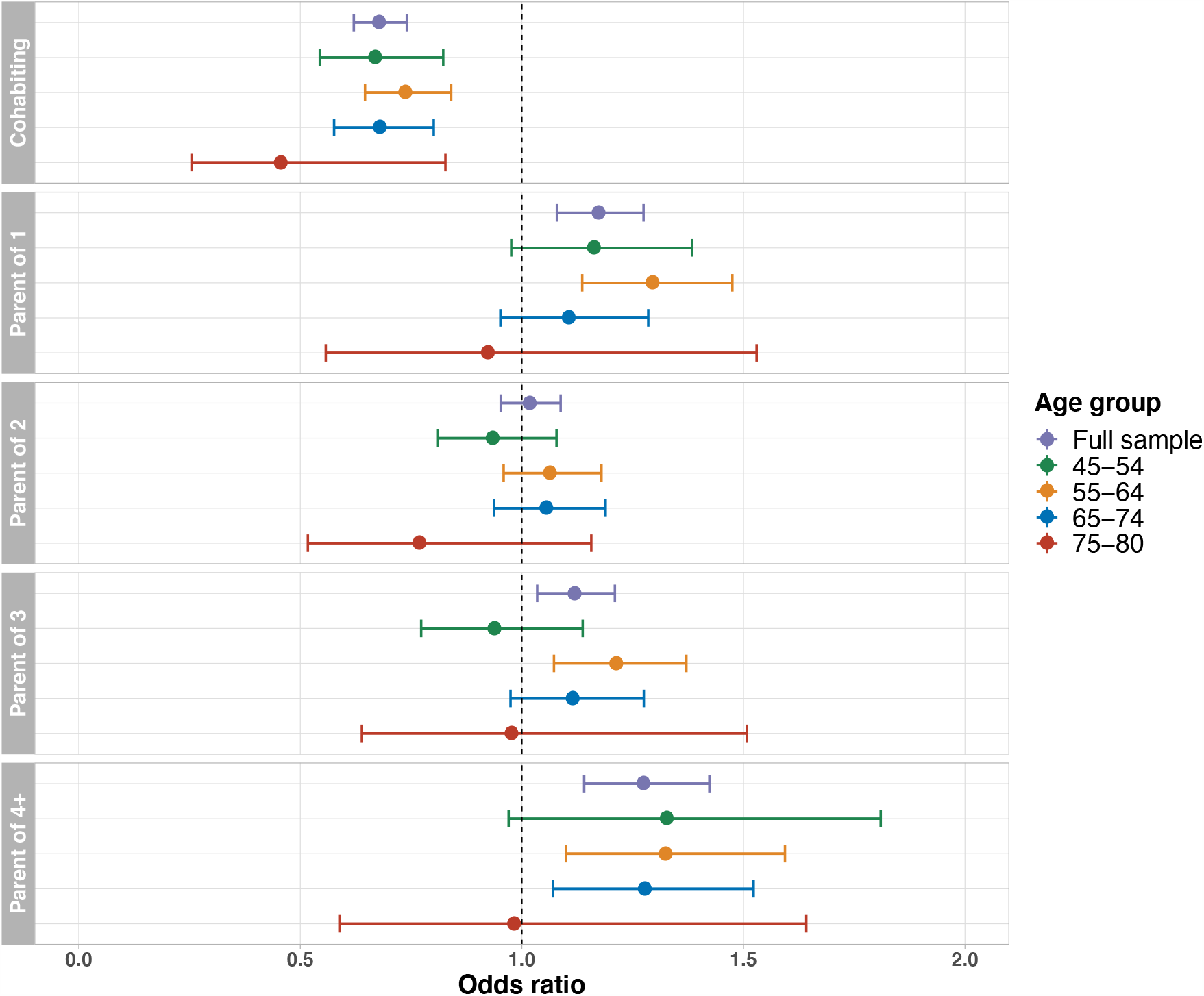
Association between family status and lifetime depression, stratified by age group. Full model including all covariates and both family status variables. Reference groups: childless; not living with a spouse/partner.

**Figure 4.**
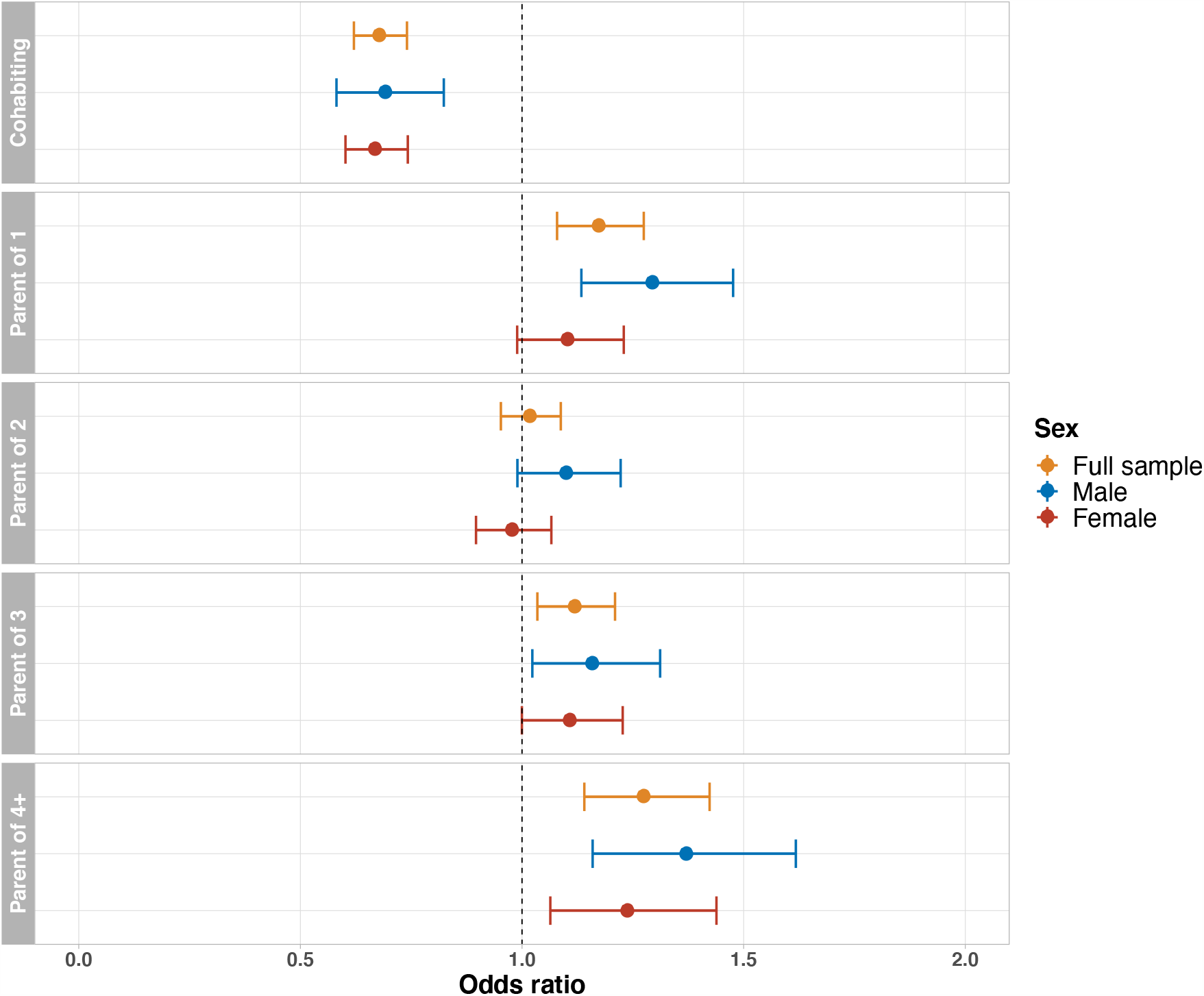
Association between family status and lifetime depression, stratified by sex. Full model including all covariates and both family status variables. Reference groups: childless; not living with a spouse/partner.

**Figure 5.**
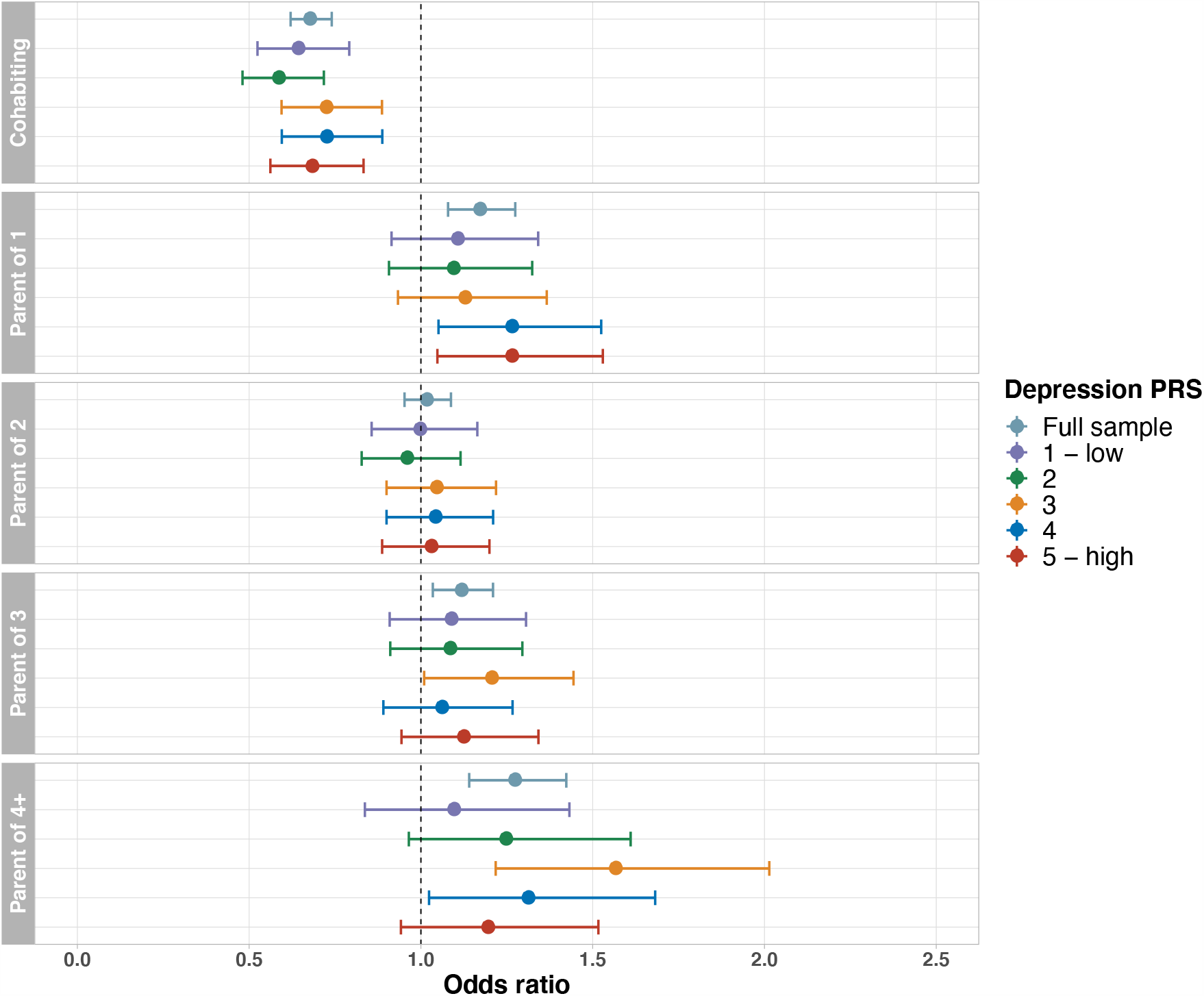
Association between family status and lifetime depression, stratified by depression polygenic risk score quintile. Full model including all covariates and both family status variables. Reference groups: childless; not living with a spouse/partner.

**Figure 6.**
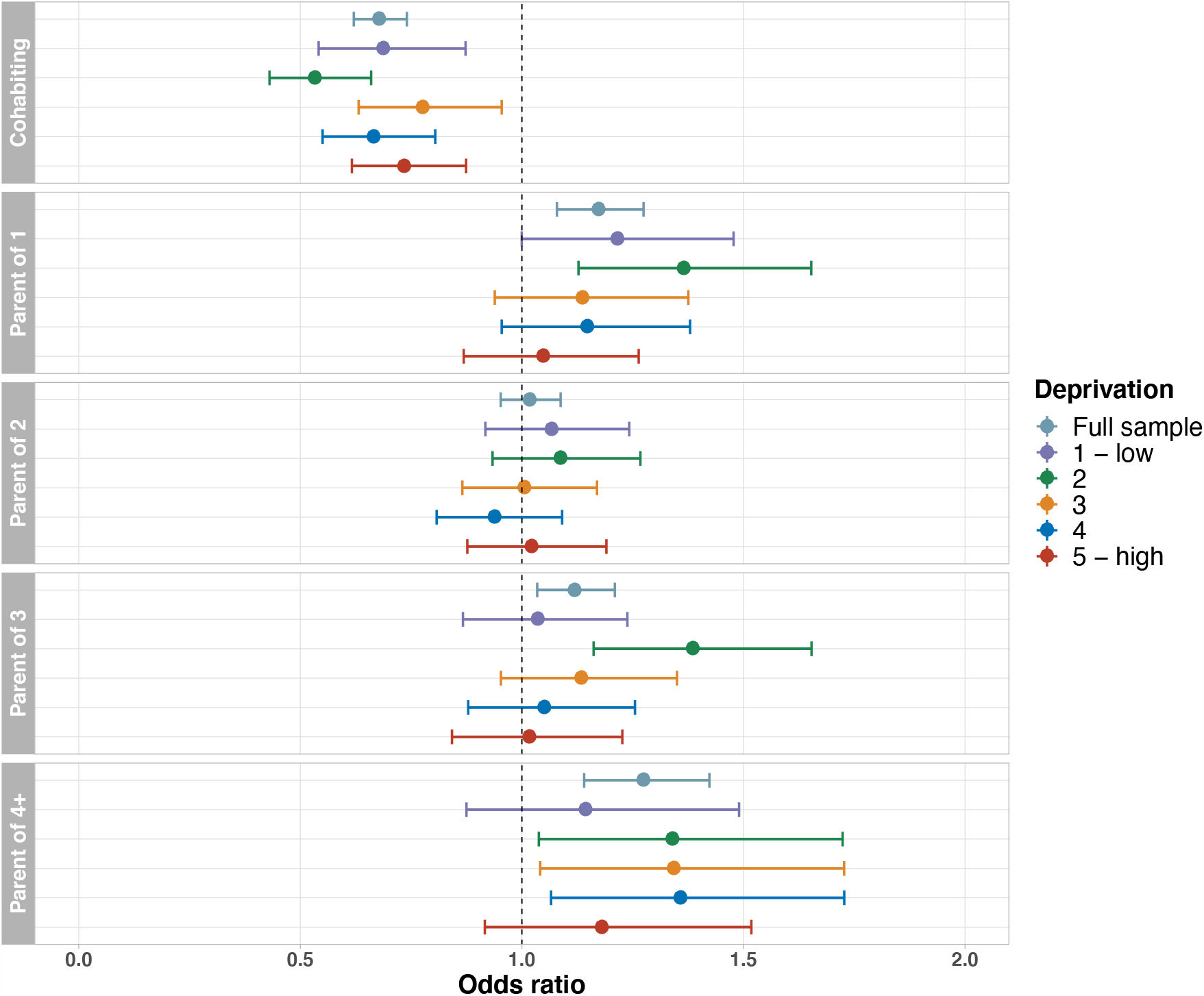
Association between family status and lifetime depression, stratified by Townsend deprivation index quintile. Full model including all covariates and both family status variables. Reference groups: childless; not living with a spouse/partner.

In the analysis in which we examined the association between number of children and lifetime depression stratified by cohabitation status, we found substantial differences between individuals living with or without a spouse or partner (Figure 7). For individuals living together with a spouse or partner, associations between number of children and lifetime depression followed a similar pattern as in the main analysis. However, amongst individuals not living with a spouse or partner, we found that parenthood was consistently associated with higher odds of lifetime depression. For example, parents of three who were not cohabiting had 122% higher odds of lifetime depression compared to childless individuals not living with a spouse or partner. For parents of two children and parent of three children there was also evidence of an interaction in the full analytical sample (both *p*_interaction_ < 0.001). However, there was no evidence of an interaction between cohabitation status and having one child or four or more children (*p*_interaction_ = 0.07 and 0.68, respectively).

**Figure 7.**
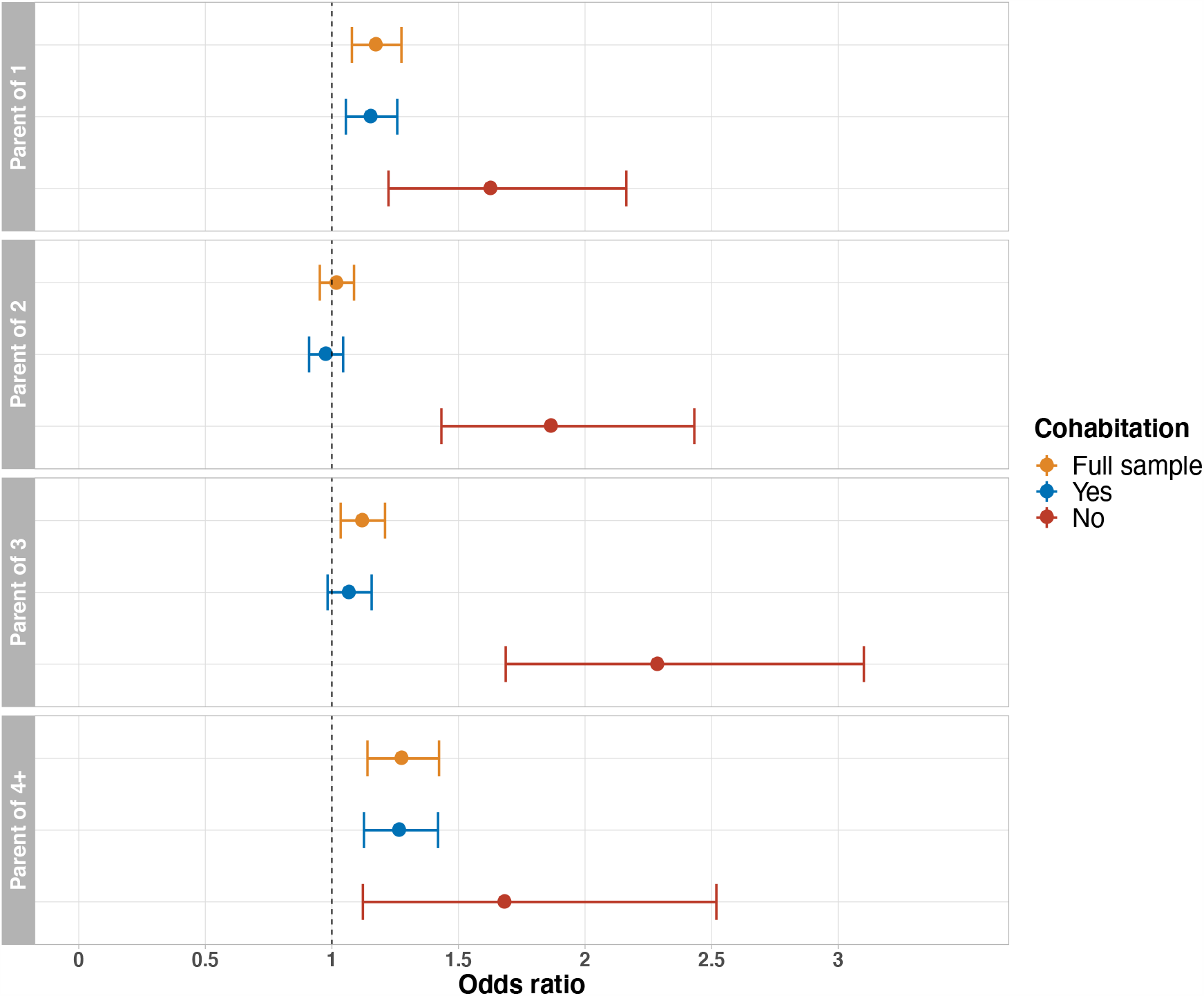
Association between parenthood and lifetime depression, stratified by cohabitation status. Full model including all covariates and number of children. Reference group: childless.

### Sensitivity analyses

Table 3 presents the results of the sensitivity analyses. Excluding women who reported postnatal depression (*n* = 1 935) from the analytical sample resulted in almost identical results for cohabitation status. However, associations between number of children and lifetime depression were attenuated, and there was no longer evidence of an association between having three children and lifetime depression (OR = 1.02, 95% CI 0.94-1.10). Examining lifetime severe depression, current depression and current severe depression yielded similar results for cohabitation status. With respect to number of children, the magnitude of associations and corresponding uncertainty estimates increased, and there were some inconsistencies in findings for current and current severe depression. There was no evidence that having two children was associated with depression across the different phenotypes.

**Table 3.** Sensitivity analyses

|  | Odds ratios and 95% confidence intervals |  |  |  |  |
| --- | --- | --- | --- | --- | --- |
|  | Lifetime depression (excluding PND) | Lifetime severe depression | Current depression | Current severe depression | Lifetime depression |
|  | *n* = 50 143 | *n* = 38 990 | *n* = 40 727 | *n* = 40 551 | *n* = 52 078 |
|  | Yes: 13 953<br>No: 36 190 | Yes: 2 333<br>No: 36 657 | Yes: 896<br>No: 39 831 | Yes: 720<br>No: 39 831 | Yes: 15 421<br>No: 36 657 |
| **Cohabitation status** (living with spouse/partner) |  |  |  |  |  |
| No | Ref | Ref | Ref | Ref | Ref |
| Yes | 0.67***<br>(0.61, 0.74) | 0.60***<br>(0.50, 0.72) | 0.58***<br>(0.45, 0.76) | 0.65**<br>(0.49, 0.89) | 0.67***<br>(0.62, 0.74) |
| **Number of children** |  |  |  |  |  |
| Childless | Ref | Ref | Ref | Ref | Ref |
| Parent of 1 | 1.10*<br>(1.01, 1.20) | 1.28**<br>(1.06, 1.54) | 1.45*<br>(1.09, 1.92) | 1.55**<br>(1.13, 2.13) | 1.17***<br>(1.07, 1.27) |
| Parent of 2 | 0.94<br>(0.88, 1.01) | 1.13<br>(0.97, 1.31) | 1.10<br>(0.87, 1.40) | 1.11<br>(0.85, 1.46) | 1.01<br>(0.95, 1.08) |
| Parent of 3 | 1.02<br>(0.94, 1.10) | 1.26*<br>(1.05, 1.51) | 1.18<br>(0.88, 1.57) | 1.23<br>(0.89, 1.70) | 1.11*<br>(1.03, 1.20) |
| Parent of 4+ | 1.15*<br>(1.02, 1.28) | 1.76***<br>(1.38, 2.23) | 1.85**<br>(1.27, 2.68) | 2.10***<br>(1.39, 3.15) | 1.27***<br>(1.14, 1.42) |
| *Note:* PND = postnatal depression. All analyses conducted using the full model that included cohabitation status and number of children, adjusted for age, sex, marital separation/divorce (in the two years prior to assessment), death of spouse/partner (in the two years prior to assessment), migrant status, highest educational/professional qualification, annual gross household income, employment status, Townsend deprivation index, smoking status, alcohol use, neuroticism, long-standing illness, disability or infirmity, traumatic life events, adverse childhood experiences, participation in social/leisure activities, loneliness, ever had same-sex intercourse, lifetime number of sexual partners, body mass index, depression polygenic risk score, six ancestry-informative principal components, batch number and assessment centre. *** $p < 0.001$, ** $p < 0.01$, * $p < 0.05$ (corrected for multiple testing at 5% false discovery rate). |  |  |  |  |  |

### Mendelian randomisation

Our findings show that the genetic instruments for number of children were associated with increased odds of lifetime depression (OR = 1.98, 95% CI 1.23-3.20, *p* = 0.005). The genetic instruments for cohabitation status were associated with approximately 23% lower odds of lifetime depression, although the effect was not statistically significant (OR = 0.77, 95% CI 0.36-1.65, *p* = 0.498). We did not find evidence that the genetic instruments for lifetime depression were associated with number of children (OR = 1.004, 95% CI 0.99-1.02, *p* = 0.581) or cohabitation status (OR = 1.006, 95% CI 0.99-1.02, *p* = 0.428). These findings are presented in Figure 8.

**Figure 8.**
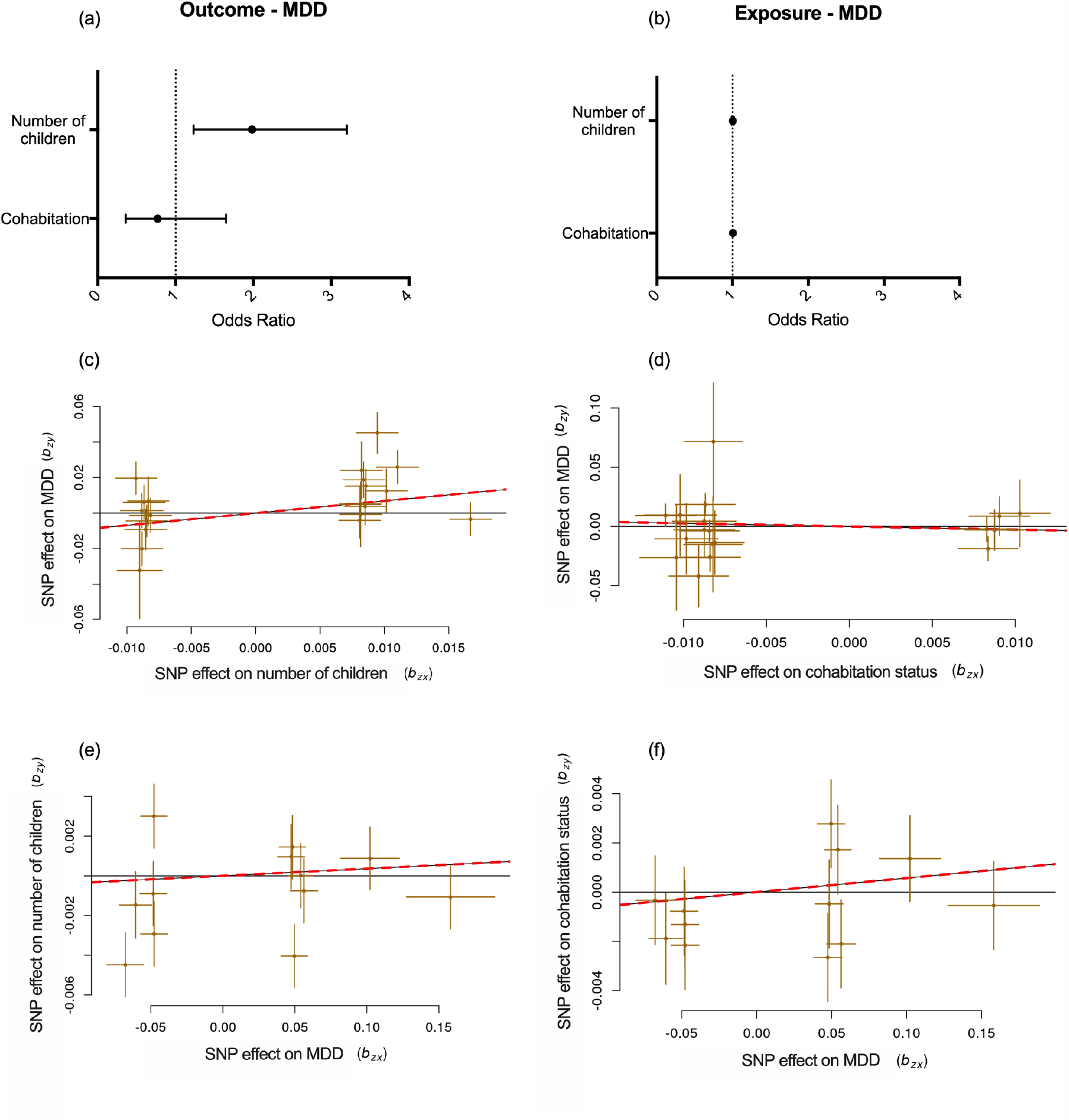
MDD = major depressive disorder; SNP = single nucleotide polymorphism. Panel (a) shows the combined GSMR estimated effect sizes (odds ratio ± 95% confidence interval) of family status on lifetime depression. Panel (b) shows the combined GSMR estimated effect sizes of lifetime depression on family status. Panel (c) shows the scatterplot of SNP potential effects on number of children vs lifetime depression. Panel (d) shows the scatterplot of SNP potential effects on cohabitation status vs lifetime depression. Panel (e) shows the scatterplot of SNP potential effects on lifetime depression vs number of children. Panel (f) shows the scatterplot of SNP potential effects on lifetime depression vs cohabitation status. For panels (c) – (f), the slope of the line indicates the estimated MR effect. Data in these panels are expressed as raw *β* values with standard errors.

The GSMR results were confirmed using the IVW and MR-RAPS methods, which showed that the genetic instruments for number of children were associated with an increased odds of lifetime depression (IVW: OR = 1.98, 95% CI 1.03-3.80, *p* = 0.039; MR-RAPS: OR = 2.05, 95% CI 1.05-4.01, *p* = 0.039). There was, however, no evidence of any other significant associations, similar to what we found using GSMR. MR-Egger provided no evidence of significant associations. See Supplement 9 for a full table of these results.

## Discussion

In the present study we showed that living with a spouse or partner was associated with substantially lower odds of depression. This association remained after extensive adjustment for potential confounders and was robust to sensitivity analyses. Additional adjustment for number of children had little impact on the association between cohabitation status and depression.

Most individuals in the present study who experienced marital separation or divorce or the death of a spouse or partner in the two years prior to the baseline assessment were not cohabiting, and this accounted for part of the association between cohabitation status and depression. Additionally, 74% of individuals not living with a spouse or partner were female, who typically report higher rates of depression, and adjusting for sex slightly attenuated the association between cohabitation status and depression. Low annual household income, adverse childhood experiences and ever having had same-sex intercourse were more frequent amongst participants not living with a spouse or partner, which is line with previous studies (57,58). When we adjusted for these and other potential confounders, we still found a substantial association between cohabitation status and depression.

We found similar associations when we examined lifetime severe depression, current depression and current severe depression, and the results were consistent across age groups, the sexes, levels of neighbourhood deprivation and depression PRS quintiles. These findings suggest that living with a spouse or partner was associated with lower odds of depression, irrespective of the social, economic or genetic characteristics that we adjusted for. Of particular interest is the absence of any interaction with sex. Previous findings from genetic studies suggested a beneficial effect of marriage exclusively for women (26). There have also been claims that marriage is only beneficial for men, and harmful for women’s happiness and wellbeing (59). Our findings do not support such views, since the association between cohabitation status and depression was the same in both sexes.

The direction of causation was explored in the MR analyses. The results suggest that living with a spouse or partner is causally linked with lower odds of lifetime depression. The effect was not statistically significant, however this may be due to reduced power, given the limited number of genetic instruments that were used. Large-scale genome-wide association studies are required in order to identify more SNPs related to family status. The MR analyses did not provide evidence in support of a causal effect of lifetime depression on family status. These results, although not conclusive, give more credence to the hypothesis that partnership is a protective factor against depression (26–28), as opposed to that of depression-prone individuals tending not to form relationships (18). The finding that the association between cohabitation status and depression was consistent across depression PRS quintiles suggests that living with a spouse or partner is associated with reduced odds of depression, regardless of genetic predisposition to depression.

It has been suggested that married individuals or those in partnership are less likely to be exposed to unpleasant experiences which may induce depression (25). In the present study we found that the association between cohabitation status and depression remained after controlling for traumatic life events and other risk factors, suggesting that this association could not be fully accounted for by married or cohabitating individuals having fewer adverse experiences. An additional explanation, which is more consistent with our results, is that living with a spouse or partner provides a source of intimacy and confidante support which might reduce the risk of depression (25). It is worth noting that we also adjusted for self-reported loneliness. Therefore, we can infer that the association between cohabitation status and depression is not driven only by the absence of loneliness in cohabiting individuals, but that there is something unique about partnership per se, which is associated with reduced rates of depression. This conclusion is also supported by the fact that individuals who did not live with a spouse or partner were not necessarily living alone but might have been cohabiting with other relatives or unrelated individuals.

We examined the broad category of “living with a partner or spouse” as no information on marital status was available in the UK Biobank. This is both a strength and potential limitation. We provide evidence that living with a spouse or partner per se, and not necessarily any legal or financial conditions associated with marriage, was associated with lower odds of depression. However, other research has suggested that there are differences in the effect of partnership on depression between married and non-married cohabiting couples (31), and we could not explore this in the current study.

Associations between number of children and depression were less consistent. When we included only number of children in the model, having one child was associated with slightly higher odds of depression, while having two children or having three children was associated with lower odds of depression. However, the associations between having two or three children and depression were not consistent when we accounted for other variables in the adjusted models. It is therefore likely that potential confounding might explain some of the associations observed in the unadjusted model. For example, having a college degree is associated with higher rates of depression (47) and having fewer children (60), therefore educational attainment is an obvious candidate for confounding the association.

Our findings provide some support for the association between number of children and depression that was recently observed in genetic studies (61). Having four or more children was associated with a 27% increase in the odds of reporting lifetime depression in the fully adjusted model, possibly reflecting the stress and economic strain that might result from having multiple children. The magnitude of association attenuated when we excluded women who reported depression that was possibly related to childbirth, suggesting that postnatal depression is an important factor contributing to the association between number of children and depression. The MR analyses provide evidence of a causal effect of number of children on lifetime depression.

In the stratified analyses, we observed that the association between number of children and lifetime depression was much stronger in non-cohabiting individuals, which is in line with previous findings (33). Financial and emotional challenges, social stigma and lack of social support may all be involved in this association. However, given that the sample included middle-aged and older adults, it is not certain that those participants are all single parents as some of them might have only become single after their children had grown up.

### Strengths and limitations

A major strength of the present study was the large sample size (> 50 000 participants), which allowed for high precision in the estimation of associations and for modelling interactions. Moreover, the wealth of information available through the UK Biobank allowed us to adjust for many of the factors that have previously been linked to family status and depression. Of particular importance, in our view, is that we also included a polygenic risk score to adjust for genetic susceptibility to depression. This study illustrates how social science can incorporate genetic data, not only in the form of twin studies, which necessarily have limited sample size, but by using molecular data available for tens of thousands of individuals.

Several potential limitations need to be considered in evaluating the present study. The UK Biobank, and the MHQ sample in particular, have been shown to not be representative of the UK general population (62). Participants were more likely to have better health and higher socioeconomic status than the average UK citizen (63). This fact is reflected in our sample, where participants disproportionally belonged to higher income, higher education and less deprived groups. In response to these concerns, UK Biobank has released a statement clarifying that, although its data cannot be used to provide representative disease prevalence rates, associations between exposures and outcomes are nonetheless widely generalisable, although there might be some differences in the magnitude of associations(63). Furthermore, the UK Biobank includes middle aged and older adults for whom perceptions of family life may be different from those of younger people. Raising young children might also be different from being a parent of independent adults, in terms of stress and responsibilities. Regarding number of children, we only included biological children and did not take adoptees into account. As such the present findings might not generalise to adoptive parents. The depression PRS that we used currently explains only a small percentage of the variance in depression, which might explain the absence of any interaction with family status in our study. Finally, our findings might not generalise to non-European ancestry groups.

Several limitations pertaining to the MR analyses should be noted. First, we lowered the *p*-value thresholds for all GWAS in order to obtain enough SNP instruments to perform MR. This may introduce false positives and type 1 error. However, we also performed GSMR using only those genetic instruments for number of children that were significant above the standard *p* < 5 × 10^−8^ threshold (6 SNPs in total) and obtained similar results. Nonetheless, we could not do the same for the cohabitation status and lifetime depression GWAS and thus there is still the potential for a type 1 error. As such, these results should be treated as exploratory rather than conclusive. Second, all three GWAS in this study were carried out on complex traits and although the lifetime depression GWAS phenotype was from clinical cohorts, concerns remain about the validity of associations derived from such analyses. However, this issue is difficult to overcome in genetics when studying complex traits.

### Implications and future directions

This is to the best of our knowledge the first study to provide evidence of an association between of family status and depression in a large population-based study with extensive adjustment for both non-genetic and genetic factors. At a time in which the number of singles has peaked and the number of marriages is at an all-time low in the UK (56), our findings indicate that pair bonding is beneficial to our mental health. Although family welfare occasionally makes it to public discourse, aspects of mental health are rarely discussed. Certain potential policy recommendations stem from this work. The increased burden on mental health faced by single parents and parents of many children highlights their need for increased support.

Future studies should aim to examine our findings using official data on marital status and cohabitation status, and to extend the scope of our research to include both biological and adopted children. Moreover, further research is needed in younger cohorts. It would be particularly useful to adopt a longitudinal design, in order to infer whether depressive symptoms predate and select people into or out of partnership and parenthood, or whether differences in odds of depression become apparent after relationship formation.

## Data Availability

The data used in the present study are available to all bona fide researchers for health-related research that is in the public interest, subject to an application process and approval criteria. Study materials are publicly available online at http://www.ukbiobank.ac.uk. Data access permission for the present study has been granted under UK Biobank application 45514.

